# Probing a neural unreliability account of auditory sensory processing atypicalities in Rett Syndrome

**DOI:** 10.1101/2024.01.25.24301723

**Authors:** Tufikameni Brima, Shlomit Beker, Kevin D. Prinsloo, John S. Butler, Aleksandra Djukic, Edward G. Freedman, Sophie Molholm, John J. Foxe

**Author notes:** Equal contributions (Tufikameni Brima, Shlomit Beker).

## Abstract

**Background:** In the search for objective tools to quantify neural function in Rett Syndrome (RTT), which are crucial in the evaluation of therapeutic efficacy in clinical trials, recordings of sensory-perceptual functioning using event-related potential (ERP) approaches have emerged as potentially powerful tools. Considerable work points to highly anomalous auditory evoked potentials (AEPs) in RTT. However, an assumption of the typical signal-averaging method used to derive these measures is “stationarity” of the underlying responses – i.e. neural responses to each input are highly stereotyped. An alternate possibility is that responses to repeated stimuli are highly variable in RTT. If so, this will significantly impact the validity of assumptions about underlying neural dysfunction, and likely lead to overestimation of underlying neuropathology. To assess this possibility, analyses at the single-trial level assessing signal-to-noise ratios (SNR), inter-trial variability (ITV) and inter-trial phase coherence (ITPC) are necessary.

**Methods:** AEPs were recorded to simple 100Hz tones from 18 RTT and 27 age-matched controls (Ages: 6-22 years). We applied standard AEP averaging, as well as measures of neuronal reliability at the single-trial level (i.e. SNR, ITV, ITPC). To separate signal-carrying components from non-neural noise sources, we also applied a denoising source separation (DSS) algorithm and then repeated the reliability measures.

**Results:** Substantially increased ITV, lower SNRs, and reduced ITPC were observed in auditory responses of RTT participants, supporting a “neural unreliability” account. Application of the DSS technique made it clear that non-neural noise sources contribute to overestimation of the extent of processing deficits in RTT. Post-DSS, ITV measures were substantially reduced, so much so that pre-DSS ITV differences between RTT and TD populations were no longer detected. In the case of SNR and ITPC, DSS substantially improved these estimates in the RTT population, but robust differences between RTT and TD were still fully evident.

**Conclusions:** To accurately represent the degree of neural dysfunction in RTT using the ERP technique, a consideration of response reliability at the single-trial level is highly advised. Non-neural sources of noise lead to overestimation of the degree of pathological processing in RTT, and denoising source separation techniques during signal processing substantially ameliorate this issue.

## INTRODUCTION

Rett Syndrome (RTT), an X-linked monogenic disorder caused by *de novo* mutations in the Methyl-CpG-binding protein 2 gene (*MeCP2*), is associated with severe intellectual disability in female children (Amir, Van den Veyver et al. 1999) (Naidu 1997). Classical RTT begins with early onset neurodevelopmental regression, typically detected between 6 to 18 months of age, that results in progressive loss of previously acquired speech and motor skills (Hagberg 2002). The inability to verbalize, a feature in the vast majority of these children, substantially impedes objective clinical assessments of their perceptual and cognitive functioning since conventional cognitive evaluations rely heavily on overt verbal or gestural responses (Berger-Sweeney 2011). As such, primary outcome measures in RTT are generally based on clinical judgement. As a consequence, there is limited knowledge about the perceptual and cognitive capabilities of these individuals across the progressive clinical stages of RTT (Demeter 2000) (Percy, Neul et al. 2010). The lack of objective assessment tools adversely impacts both clinical evaluation and the measurement of therapeutic efficacy during intervention trials. It is therefore imperative for the field to identify quantitative measures of neural function that can be objectively measured and longitudinally monitored to capture more subtle changes in neurological function (Saby, Peters et al. 2020) (Demeter 2000), ideally without the need for active task participation on the part of these individuals given the typical severity of the phenotype. Developing such measures would provide surrogate biomarkers of disease severity and potentially provide precise measurement of target engagement and longitudinal evaluations of treatment effects during clinical trials (Saby, Peters et al. 2020) (Demeter 2000).

To this end, a number of research groups have now deployed electroencephalographic (EEG) recordings as a means to directly measure brain function in neurodevelopmental disorders (Ortiz, Martinez-Murcia et al. 2020) (Rossion, Retter et al. 2020) (Banaschewski and Brandeis 2007) (Sueyoshi and Sumiyoshi 2018) (Black, Chen et al. 2017) (Butler, Molholm et al. 2017) (Knight, Oakes et al. 2020) (Francisco, Berruti et al. 2021). EEG provides an easy-to-deploy method to assay neurodevelopmental regression in the absence of overt behavioral responses from participants (e.g. (Demeter 2000) (Banaschewski and Brandeis 2007) (Shahaf, Yariv et al. 2017) (Ortiz, Martinez-Murcia et al. 2020)). The millisecond-precision of this tool is ideal for assessment of dynamic brain function and can be used to determine the processing level at which information flow is breaking down (Foxe and Simpson 2002, Muller, Vetsch et al. 2020) (Shahaf, Yariv et al. 2017). This is achieved by assaying the latencies and amplitudes of well-characterized event-related potential (ERP) components, which have stereotypical topology and temporal dynamics in neurotypical populations, and have been well characterized in thousands of papers over the past 60 years (Chope, Metz-Lutz et al. 1994) (Luck 2014) (Simpson, Pflieger et al. 1995) (Martin, Barajas et al. 1988) (Sutton, Tueting et al. 1967) (Sutton, Braren et al. 1965) (Ritter and Vaughan 1969). A high degree of test-retest reliability is also a feature of this method, making it ideal for longitudinal monitoring of intervention trials (Kileny and Kripal 1987) (Malcolm, Foxe et al. 2019) (Beker, Foxe et al. 2021). However, a central assumption of this methodology is stationarity of response – that is, that when a stimulus is presented repeatedly to a participant, the neural response on each iteration (or trial) is assumed to be essentially identical, whereby the simple process of signal-averaging across trials will reveal this stationary canonical response because temporally random background activity (noise) will be eliminated through the averaging procedure (Luck 2014, Helfrich and Knight 2019) (Ritter and Vaughan 1969). While this is perhaps not an unreasonable assumption in studies of neurotypical individuals, it may not be fully justified to assume that near-perfect stationarity is a feature of sensory perceptual processing in neurodevelopmental and neuropsychiatric conditions. For example, a number of researchers have proposed that the neural response in autism spectrum disorder (ASD) may be more variable, or unreliable, on a trial-to-trial basis (Haigh, Brosseau et al. 2022) (Milne 2011) (Haigh, Heeger et al. 2015) (but see (Butler, Molholm et al. 2017) (Kovarski, Malvy et al. 2019) (Dwyer, Vukusic et al. 2022)). That is, it could be the case that an evoked response is produced to each stimulus iteration in these conditions, but that from trial to trial, this evoked response might vary in the latencies and amplitudes of the canonical components. In such a situation, signal averaging will have the same impact on these “signals” as it does on the background noise – that is, they will tend to reduce towards zero.

To date, most ERP studies in Rett have shown highly disordered sensory responses, both in audition (Foxe, Burke et al. 2016) (Key, Jones et al. 2019) (Saby, Benke et al. 2021) (Sysoeva, Molholm et al. 2020) (Brima, Molholm et al. 2019) (Peters, Katzenstein et al. 2017) (Badr, Witt-Engerstrom et al. 1987) and vision (Millichap 2015) (LeBlanc, DeGregorio et al. 2015) (Stauder, Smeets et al. 2006) (Saunders, McCulloch et al. 1995) (Kalmanchey 1990) (Bader, Witt-Engerstrom et al. 1989), but to our knowledge, all of these previous studies, including those from our own research group, rely on the standard signal-averaging approach. Here, we are interested in determining whether a higher degree of variability, both at the individual participant and at the group level, might be a factor in the reduced and delayed ERP responses typically reported in RTT. This is important, because it can have significant implications for the use of the standard ERP as a neuromarker in clinical trials, and it is plausible that functionality at the individual participant level is being obscured by the averaging technique. This is also the case at the group level, where idiosyncratic ERP morphologies and timings at the individual level, when averaged together across the group, could potentially give the impression that the group has a much greater overall deficit than is actually the case.

We set out to test what we have termed the “unreliability account” by measuring the coherence of the auditory evoked potential (AEP) in both RTT and neurotypical (NT) age-matched controls at the single trial level, with an eye to more deeply characterizing potential auditory functionality in RTT. We recorded the AEP from a 72-channel montage in response to simple 1 kHz pure tones at three different stimulus-onset-asynchronies (SOA’s: 450, 900 and 1800 ms, see (Brima, Molholm et al. 2019), in which standard group analyses of the averaged ERP focused on the mismatch negativity (MMN) response is reported for the same dataset). The relatively large numbers of trials presented to each participant (∼500 per condition on average, see Table S2) allowed for in-depth analysis at the single trial level, which is necessary for measures of inter-trial (i.e. intra-participant) variability with high statistical power. To measure inter-trial reliability of the auditory responses, we applied a number of relevant approaches, calculating inter-trial-variability (ITV), signal-to-noise estimates (SNR), and inter-trail phase coherence (ITPC) at the individual participant level. We additionally sought to better understand the inter-participant variability that may derive from combining participants across various stages of disease severity by comparing homogeneity of the AEP between the groups.

Another consideration when making EEG/ERP recordings in clinical populations is baseline differences in non-neural sources of noise, such as those produced by muscle or movement artifacts (Goncharova, McFarland et al. 2003), which can also serve to reduce the reliability of estimations of neural activity and potentially lead to overestimation of inter-group differences. To this end, we applied data denoising source separation (DSS) to separate temporally coupled signal carrying components from temporally decoupled activity (Särelä, Valpola et al. 2005) (de Cheveigne and Simon 2008) (Granados Barbero, De Vos et al. 2021), and compared all of the above measures post-compared to pre-DSS signal derivation.

## MATERIALS AND METHODS

### Participants

Data were analyzed from 25 females with confirmed *MECP2* mutations and 30 typically developing controls (TD) (20 females and 10 males). Participants with RTT were recruited through the Rett Syndrome Center of Montefiore Children’s Hospital in the Bronx, NY, while TD participants were recruited from the local community. Seven datasets from the RTT group and three from the TD group were excluded from further analysis due to noisy EEG data that resulted in less than 20% accepted trials per condition. The final sample contained 17 females with RTT (mean age: 12.6±4.8, range 6-22) and 24 TDs (15 females and 9 males) (12.45±4.9, range 6-26). There was no significant difference in age between the RTT and TD group (t (41) = 0.12, p = 0.9).

All participants with RTT underwent genetic testing and phenotypic assessment accompanied by detailed medical history questionnaires completed by their caregivers. Symptom severity in RTT was measured using the Rett Syndrome Severity Scale (RSSS) which is the primary scale used by the Rett Syndrome Center of Montefiore Children’s Hospital (Kaufmann, Tierney et al. 2012) (Kaufmann, Tierney et al. 2012, Neul, Glaze et al. 2015). This clinician-rated scale represents an aggregate measure of the severity of clinical symptoms, including motor function, seizures, autonomic function, ambulation, eye contact, and communication (Neul, Glaze et al. 2015). The RSSS score in the current RTT group ranged between 5 and 15 (Mean ± SD = 10.94±2.8), with higher scores indicating more severe disease. For reference, composite scores in the range of 0– 7 are considered to correspond to a mild phenotype, 8-14 to a moderate phenotype, and 15-21 to severe features (Kaufmann, Tierney et al. 2012).

TDs were excluded if they had a family history of a neurodevelopmental disorder or any neurological/psychiatric disorders. All individuals in the TD group passed a hearing screen on the day of EEG testing. A limitation of the current study is that hearing acuity could not be similarly assessed in participants with RTT. However, in all cases, parents reported that the children with RTT could hear, and this was confirmed by clinical observation. Furthermore, participants with RTT were excluded if they had evidence of ear infection on the day of EEG acquisition. Tympanometry was performed on all participants to rule out middle-ear involvement, and Type-A tympanograms were observed in all cases. Clinical demographic information, including RSSS severity scores, ages of onset and regression, and medication of all participants, are listed in supplementary materials (Table S1; *Clinical Demographics*). There were no differences in age-range or RSSS scores between the seven excluded RTT datasets, and those included in the final analysis.

All aspects of the research conformed to the tenets of the Declaration of Helsinki. The institutional review boards of the University of Rochester and the Albert Einstein College of Medicine approved this study. Written informed consent was obtained from parents or legal guardians, and where possible, informed assent from the participants was obtained. Participants were compensated at a rate of $15/hour for their time.

### Experimental Design, Procedure and Stimuli

Experimental design, procedures and stimuli were identical to those described in an earlier report from this dataset (Brima, Molholm et al. 2019) and have been purposefully deployed in a number of other rare disease populations to allow for comparisons across phenotypes (Francisco, Berruti et al. 2021, Brima, Freedman et al. 2024) (Francisco, Foxe et al. 2020) (Francisco, Foxe et al. 2020); See Figure S1 for a paradigm schematics). We presented a simple auditory mismatch-negativity (MMN) paradigm while recording high-density EEG (72 channels). All participants sat in a sound-attenuated and electrically shielded booth (Industrial Acoustics Company, Bronx, New York) on a caregiver’s lap or in a chair/wheelchair. They watched a muted movie of their choice on a laptop (Dell Latitude E640) while passively listening to auditory stimuli presented at an intensity of 75 dB SPL using a pair of Etymotic insert earphones (Etymotic Research, Inc., Elk Grove Village, IL, USA). The MMN paradigm consisted of regularly (85%) occurring standard tones that were randomly (15%) interspersed with deviant tones, with the constraint that two deviant tones never occurred in succession. These tones had a frequency of 1000 Hz with a rise and fall time of 10 ms. Standard tones had duration of 100 ms while deviant tones were 180 ms in duration. The responses to the deviant tones were reported in our earlier paper which concentrated on the MMN response (Brima, Molholm et al. 2019), and will not be discussed or analyzed further here. The tones were presented in three separate conditions with stimulus onset asynchronies (SOAs) of 450, 900 or 1800 ms (corresponding to 2.2, 1.1 and 0.55 Hz, respectively). These SOA conditions were presented in separate blocks, with each block consisting of 500, 250 or 125 trials respectively (Fig S1A). Participants were presented with 14 blocks altogether (2×450ms, 4×900ms and 8×1800ms), resulting in 1000 trials per condition. Only the responses to the standard 100 ms tones are analyzed here.

### EEG Acquisition

A Biosemi ActiveTwo (Bio Semi B.V., Amsterdam, Netherlands) 72-electrode array was used to record continuous EEG signals. The setup includes an analog-to digital converter, and fiber-optic pass-through to a dedicated acquisition computer (digitized at 512 Hz; DC-to-150 Hz pass-band). EEG data were referenced to an active common mode sense (CMS) electrode and a passive driven right leg (DRL) electrode.

### Data processing

EEG data were processed and analyzed offline using custom scripts that included functions from the EEGLAB Toolbox for MATLAB (the MathWorks, Natick, MA, USA) (Delorme and Makeig 2004) (Delorme and Makeig 2004) and the FieldTrip Toolbox for MATLAB (Oostenveld, Fries et al. 2011). EEG data were initially filtered using a Chebyshev Type II filter between 1Hz and 40Hz, with the following parameters: highpass filter: stopband at 0.1Hz, passband at 1Hz, attenuation: 65 dB. Lowpass filter: stopband at 40Hz, passband at 35Hz, attenuation: 65dB. Continuous EEG data were subjected to a channel rejection algorithm, which identified bad channels using measures of standard deviation and covariance with neighboring channels. Rejected channels were interpolated using the EEGLAB spherical interpolation. For all statistical analyses, data were epoched to 2 second segments: from 1 second pre-tone onset to 1 second post-tone onset. Trials with artifacts of ±150 µV were excluded from further analysis. For the remaining trials, the threshold was set at two standard deviations over the mean of the maximum values for each epoch, to exclude any remaining artifact contaminated trials. The number of accepted trials for each SOA condition and group is presented in Table S2. To maximize AEP amplitudes at the fronto-central scalp sites where analyses were carried out, data were referenced to TP7 (or TP8 if TP7 was noisy), a temporo-parietal site below the Sylvian fissure where the auditory response tends to invert relative to fronto-central sites.

### Data Analysis

All analyses were performed on data averaged from three electrodes over fronto-central scalp (FC3, FCz and FC4). A multipronged approach was taken to analyzing the data. 1) In accord with conventional ERP analyses, we tested for group level differences in the amplitude of the AEP using standard analyses of variance, for two time-windows corresponding to the two major deflections in the AEP: Average amplitudes were calculated for each participant for each group and for each SOA for the P1 (50-100 ms) and N2 (200-300 ms) timeframes. 2) Another set of analyses focused on measuring within-subject variability and comparing this across groups. For this, linear mixed effects models were applied to both regular and denoised data (as described below), with analyses on data from the N2 (200-300 ms) window, where response amplitude was greatest. Three metrics of within-subject variability were tested:

#### Signal-to-Noise Ratio (SNR)

SNR was measured across trials, for each individual in each group and for each SOA condition using a shuffling method: Signal was calculated as mean amplitude in the 200-300 ms time window, and Noise was defined as mean amplitude for the same window, with every other trial flipped in polarity (i.e., multiplied by -1) to remove the stationary response (i.e., the evoked potential).

#### Inter-trial Variability (ITV)

ITV was calculated as the mean of the deviations of the individual trials from the average AEP (standard deviations), in the 200 to 300 ms window.

#### Inter-trial phase coherence (ITPC)

To quantify the consistency of phase of the auditory response across trials, ITPC was calculated as the circular coherence of phases across trials for 2-second epochs centered on stimulus onset for each individual participant, trial, and SOA condition.

ITPC was calculated as follows (Luo and Poeppel 2007):

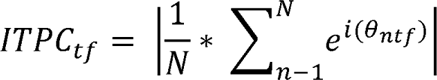

Where *θ_ntf_* is the phase at temporal bin t and frequency bin f, in trial n. Output values range between 0 (no phase coherence) to 1 (perfect phase coherence). Morlet wavelet convolution was used on the 2-second epochs. Wavelets were composed of Gaussians that ranged from 3 to 5 cycles. For visualization, ITPC is averaged across participants and presented for each group and condition, pre- and post DSS. The parameter that is used for statistical analysis is the maximal ITPC value across frequencies, calculated on 200-300ms window post stimulus onset.

#### Denoising Source Separation (DSS)

Recordings of EEG signals inherently contain both stimulus-driven responses and stimulus irrelevant responses/noise (de Cheveigne and Simon 2008, de Cheveigné and Parra 2014). In order to extract components that are directly related to auditory stimulus evoked activity, we employed dimensionality reduction through the Denoising Source Separation (DSS) algorithm (Särelä, Valpola et al. 2005) (de Cheveigne and Simon 2008). DSS decomposes multi-channel EEG recordings to extract neural response components that are consistent across trials and has been demonstrated to be effective in denoising auditory evoked activity (de Cheveigne and Simon 2008). This denoising technique is based on a blind source separation that removes stimulus-unrelated components from stimulus-related components through a spatial filter. These spatial filters are linear combinations of the sensors designed to partition data into signal carrying components of interest and non-signal carrying components (de Cheveigne and Simon 2008). In this study, DSS was performed on the 2-second-long epochs for each subject and each of the three conditions independently (presented here as *Pre-DSS* signals). After data from all channels were normalized, they were submitted to principal component analysis (PCA). This yielded a time series matrix, ordered by decreasing bias scores, that is partitioned to signal and noise components. Based on SNR calculation, it was determined that the first two DSS components contributed significantly and were optimal. These components were retained and projected back to sensor space to obtain the denoised EEG data (referred to hereafter as *Post-DSS* signals), which denote denoised auditory responses throughout this paper.

### Statistical analyses

#### Analysis of Variance (ANOVA)

In line with the standard approach for analyzing ERP components (Brima, Molholm et al. 2019) (Sysoeva, Molholm et al. 2020), we employed repeated measures ANOVA with SOA (450 , 900, and 1800 ms) as a within participant factor and Group (RTT vs. TD) as a between participant factor. This analysis was conducted on data from fronto-central electrodes (FC3, FCz and FC4), for the aforementioned time windows corresponding to the major deflections of the AEP (P1 and N2).

#### Linear mixed effects models (LME)

For subsequent analyses, to account for random and fixed effects, including differences in neuronal variability between participants, we implemented LME on the dependent measures from the different analyses. The *fitlme* MATLAB function was used. Advantages over the standard ANOVA approach have been previously detailed (Wainwright, Leatherdale et al. 2007) (Krueger and Tian 2004) (Luke 2017). Mixed-effects models account for multiple comparisons and interactions. Condition and Group were used across all models as fixed effects. Participants were treated as random factors according to the following linear-model expression: (*LME* = (*EEG_AEP_* ∼ 1 + *SOA_condition_* + *Group_RTT,Control_* + (1|*Subjects_ID*)) and for analyses involving within-subject analyses we used (*LME* = (*EEG_AEP_* ∼ 1 + *SOA_condition_* + (1|*Subjects_ID*)) where *EEG* stands for the AEP amplitude values, SOA condition corresponds to the three stimulus presentation intervals (450 ms, 900 ms, and 1800 ms), and Group corresponds to the control and RTT cohorts.

#### Wilcoxon rank test

we used non-parametric testing to assess the presence of group effects for the following measures: AEP, SNR and ITV, in both the pre-DSS and post-DSS data, separately. This was done to have a first estimation of the differences between RTT and controls in any of these measures, prior to applying the advanced *lme* models.

#### Cluster-based permutation

Cluster-based permutation statistics were used to assess significant modulations across groups and conditions and were computed as a function of channels*time (Maris and Oostenveld 2007) (Oostenveld, Fries et al. 2011). These univariate tests were performed by means of dependent samples t-tests (p<0.5 two sided), and cluster-based permutation tests (based on a minimum of 2 channels), to control for multiple comparisons. The significance of the observed cluster-level statistic (based on the t values within the cluster) was assessed by comparison to the distribution of all permutation-based cluster-level statistics. The final cluster *p* value that we report in all figures was assessed as the proportion of 2000 Monte Carlo iterations in which the cluster-level statistic was exceeded. Cluster significance was indicated by *p* values below 0.025 (two-sided cluster significance threshold).

## RESULTS

### Standard AEP analysis

In Figure 1, the standard “stationary” AEP is plotted for each individual in both the TD (left column) and RTT (right column) groups, and mean standard amplitude for the three different SOA conditions is shown in the three rows (Panels A to C) (Rousselet, Foxe et al. 2016). Panel D shows the group-averaged waveforms over-plotted for each of the SOAs. Note that the displayed AEPs represent an average of activity from the fronto-central electrode chain (FC3, FCz and FC4) as modeled in Panel E. In Panel A (the 450 ms SOA), one can readily appreciate the general morphology of the AEP and the relative consistency across individuals in the TD group (left column), with a clear P1 in the initial response (at 50-100ms; blue shaded timeframe), followed by a second smaller positive deflection (P2, ∼150 ms) and then by a longer latency negativity (between 200-300 ms: N2). The dark green trace at the bottom of Panel A shows the group-averaged TD waveform, with standard error of the mean also indicated. Despite the general consistency of the individual participant waveforms in Panel A, it can also be appreciated that even in this TD cohort, there is a high degree of inter-participant variability. This is undoubtedly enhanced by the wide age-range of our cohort, since the morphology of the AEP changes over the course of development. In the right column of Panel, A, the same over-plotting has been conducted for participants in the RTT cohort. Here, one can appreciate that the individual traces are considerably more divergent from each other, and this is reflected in the substantially reduced amplitude of the group averaged RTT waveform (red trace, bottom right of Panel A), where only a highly reduced P1 component is evident. Nonetheless, one can also appreciate that there are individuals in the RTT cohort who are producing waveforms with large amplitude positive and negative deflections that may reflect preserved auditory processing, albeit with different temporal dynamics to those seen in TD participants. Similar patterns are also evident in Panels B and C at the two slower presentation rates (900 ms SOA, panel B and 1800 ms SOA, Panel C). As the rate of presentation is slowed, the second P2 positivity emerges more clearly in the TD cohort, whereas this is not the case in the RTT cohort. At both of these slower rates, AEP responses in the TD cohort are clearly more consistent across individuals than those seen in the RTT cohort.

**Figure 1.**
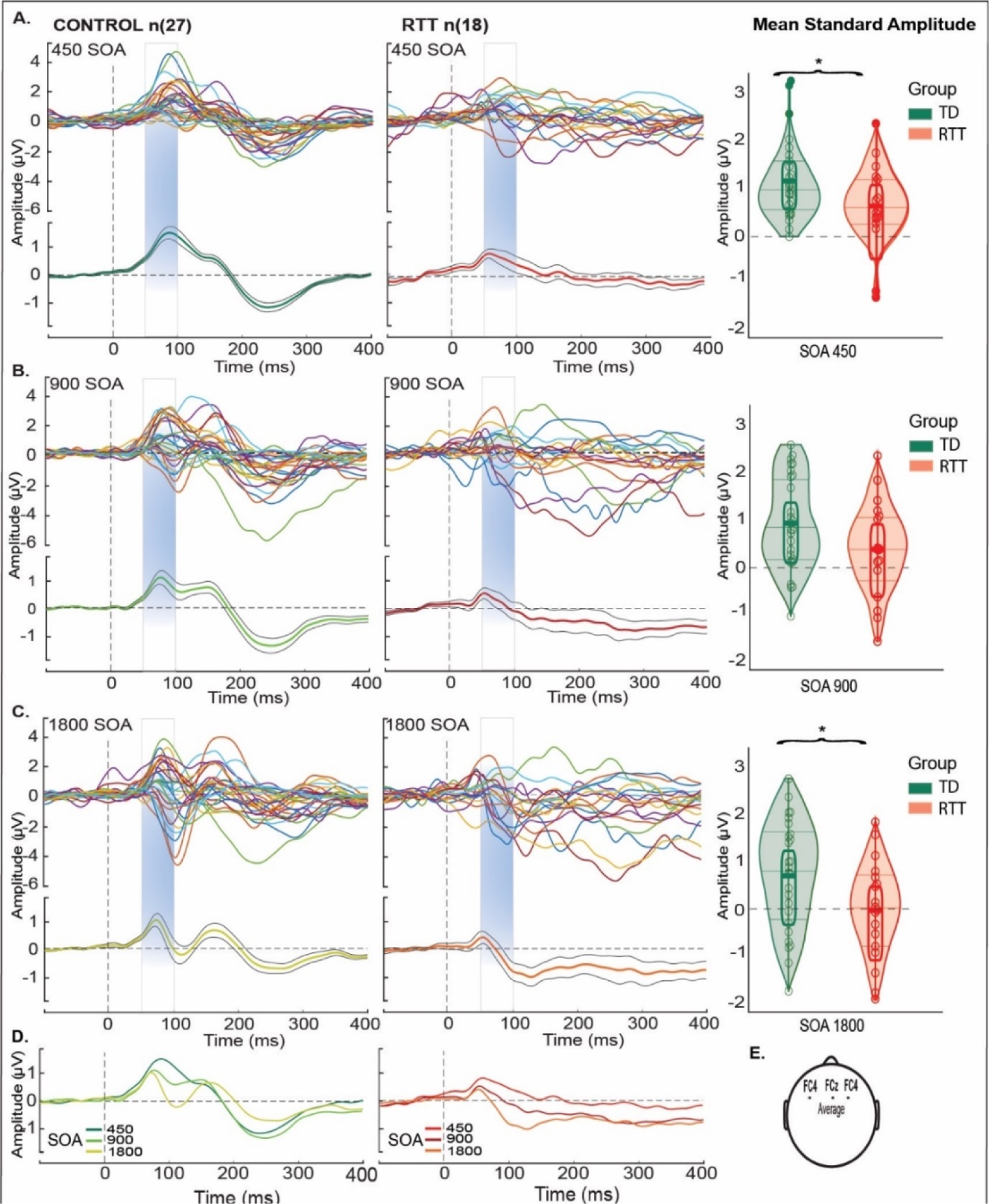
Standard Mean AEP (1 second epochs) for TDs (left) and RTT (right) over fronto-central scalp (averaged over electrodes FC3, FCz, FC4). Panels (A-C) shows colored traces representing an average of all trials in response to standard tones for each participant and their grand average AEP (green for TD and red for RTT trace with black traces – standard deviation) for all SOA conditions. TDs produced classic AEP waveforms while the RTT group exhibited atypical responses with reduced AEP amplitude across SOAs. A clear initial peak (P1) within the time period from 50 to 100 – blue shaded panels was present for all SOAs in both groups. Distribution of mean standard amplitude and quartiles are plotted at the far right in panels (A-C) for TD (green) and RTT (red) during the period of initial peak (from 50 - 100 ms) across SOAs. Significant difference between the groups is marked by asterisk (for the 450 (p=0.05), 900ms (p=0.80) and 1800ms (p=0.04) SOAs). Panel (D) shows change in AEP morphology as a function of SOA seen in the control and RTT group. Panel (E) illustrates the locations of the averaged fronto-cental scalp electors that yielded the AEPs.

For the P1, repeated measures ANOVA revealed a main effect of Group (F (2, 41) =7.4, *p* = 0.007) but not Condition (F (1, 41) = 0.71, *p* = 0.4), and no Group*Condition interaction (F(2,41) = 0.99; *p =* 0.37), indicating an attenuated AEP in RTT as compared to TD across SOA conditions. For the N2, repeated measures ANOVA revealed a main effect of Group (F(1,43) = 11.05; *p* = 0.0012) but not Condition (F(1,43) = 2.8; p = 0.06), and no Group*Condition interaction (F(1,43) = 0.26); *p =* 0.77), indicating an attenuated AEP in RTT as compared to TD across SOA conditions. All subsequent analyses are delimited to the N2 (200-300 ms) time frame.

Figure 2 shows data from four participants from each cohort randomly selected and age-matched to illustrate one of the central points of the current work. Note that panels A through D show data from children at four different age brackets (6-7, 8-9, 10-12 and 14-16 years respectively). One can readily appreciate that each of the four control participants produces highly replicable AEPs across each SOA condition – that is, there is a high degree of within-participant consistency across conditions, but also a high degree of between-participant consistency in terms of component timing and morphology, despite the relatively wide span of ages represented. In most cases, a positive deflection at about 100 ms (the P1) is followed by a broad negative deflection at about 200 ms (here referred to as N2); see grey and pink shading for timeframes used for analyses of the P1 (50-100ms) and N2 (200-300ms), respectively. In the data from participants with RTT, one can also see that AEP responses, albeit noisier (see displayed standard error of the mean (SEM) shading around the waveform traces), are highly replicable across conditions within-participant, whereas the timing and morphology of the responses are evidently not as consistent across RTT participants as they are in TDs.

**Figure 2.**
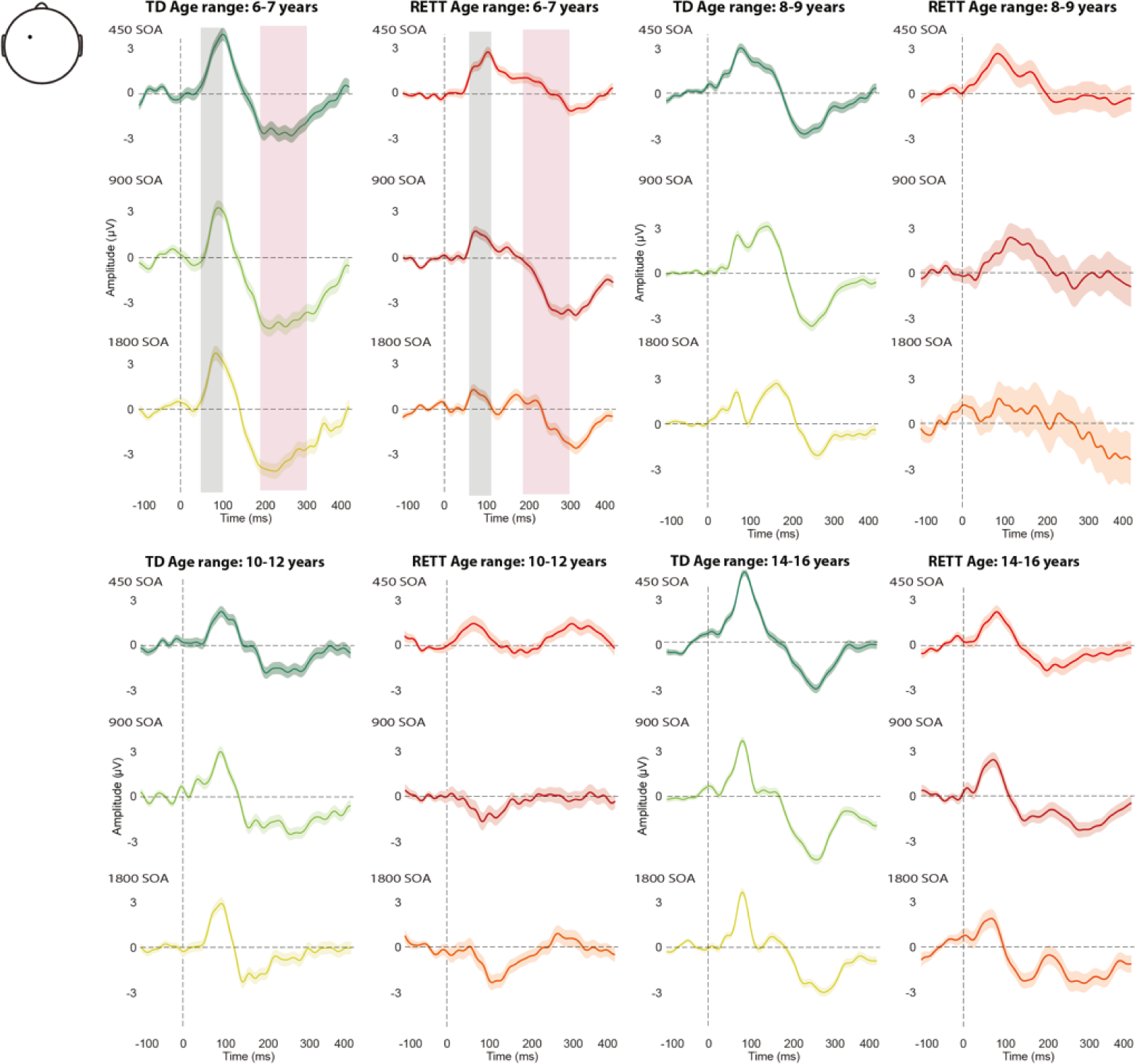
ERP waveforms in TD and RTT, for the SOA conditions. Representative Individual Participant AEP: average trials from four typically developing control participants (green shades) and four individuals with RTT (red shades) over fronto-central electrode (FC3), in 4 age ranges (6-7, 8-9, 10-12 and 14-16 years old). Gray and pink bars represent the two time windows for analysis: P1 (50-100) and N2 (200-300), respectively.

### Denoising Source Separation (DSS)

We applied the DSS technique to enhance the signal components carrying evoked activity that is reproducible across trials, with an eye to enhancing the signal to noise ratio of the AEP in the RETT data. DSS achieves this by accentuating signals which are consistent across trials while suppressing noise-like components that are independent of stimulus timing (Särelä, Valpola et al. 2005) (de Cheveigne and Simon 2008).

Figure 3 displays ERP trials pre- and post-DSS, for each group and SOA condition. Using the DSS denoised data, we replicated the initial AEP analyses, over the same fixed N2 time-window (200-300ms) and averaged across the same channels (FC3, Fz, FC4), to further explore any difference that might be revealed using DSS (i.e., Post-DSS data). AEP values for N2 pre and post DSS are shown in Figure 4. Pre-DSS, TD individuals had significantly greater AEP N2 amplitudes compared to RTT (*RTT: Mean ± sem = -0.17 ± 0.22; TD: Mean ± sem = -0.69 ± 0.44; Wilcoxon rank test statistic = 3781; p = 4.58e-04)*. Post-DSS, this difference in AEP N2 was no longer detected (*RTT: Mean ± sem = -0.18 ± 0.19; TD: Mean ± sem = -0.40 ± 0.15; Wilcoxon rank test statistic = 4093; p = 0.057)*).

**Figure 3.**
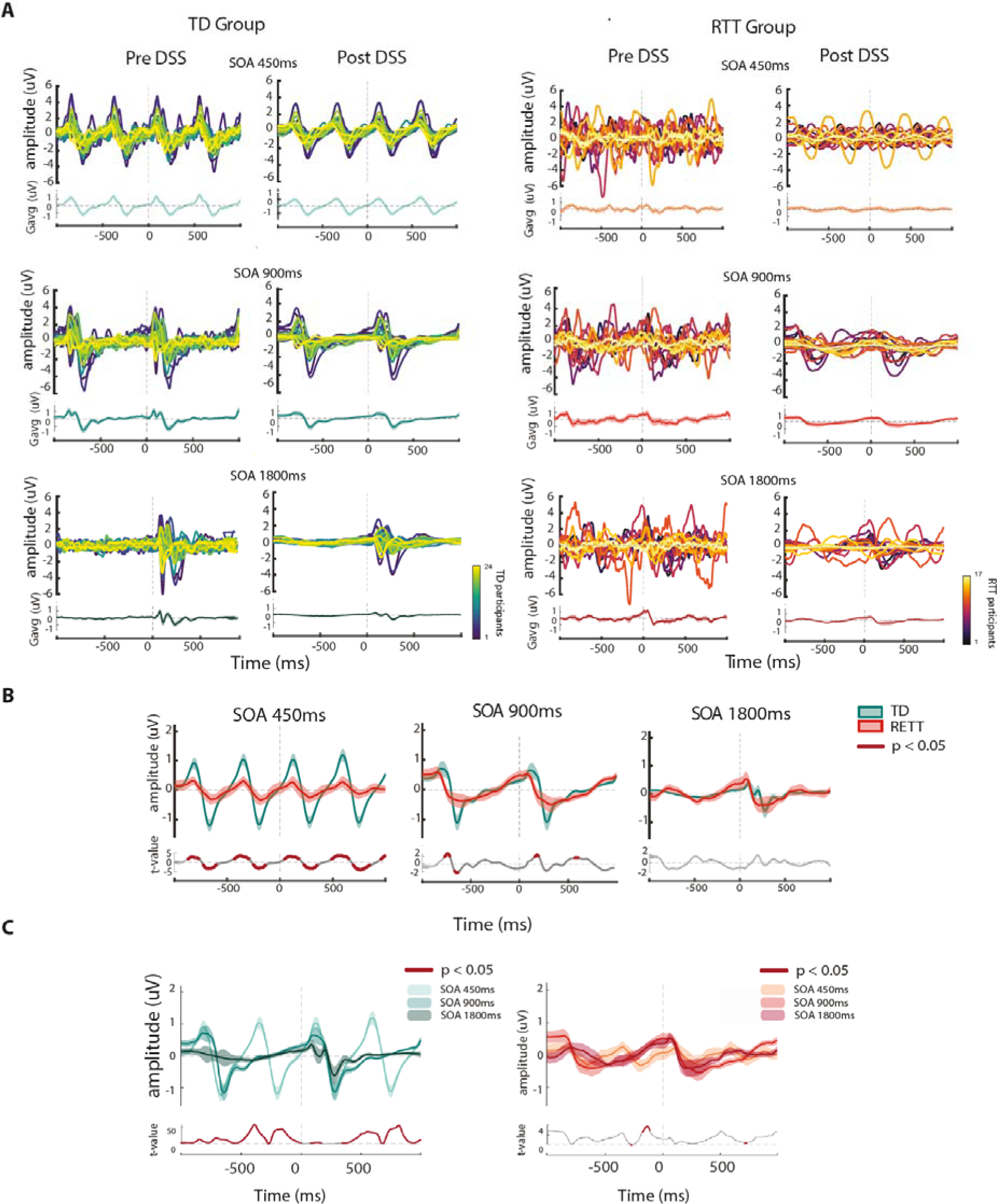
ERP waveforms in TD and RTT, pre-and post DSS. A. participant waveforms in response to standard tones overlaid (top panels) and averaged (bottom panels) across individuals, shown for each Group and for each SOA condition, pre- and post denoise source separation (DSS). B. Group averaged ERP signals (mean ± SEM in shaded lines) compared N2 between TD and RTT, for each SOA condition. Lower panels show cluster permutation statistics between groups (thick red line indicates significant temporal regions, *p* < 0.05). C. Same as B, however a comparison is presented between averaged ERPs for all SOA conditions in each of the groups.

**Figure 4:**
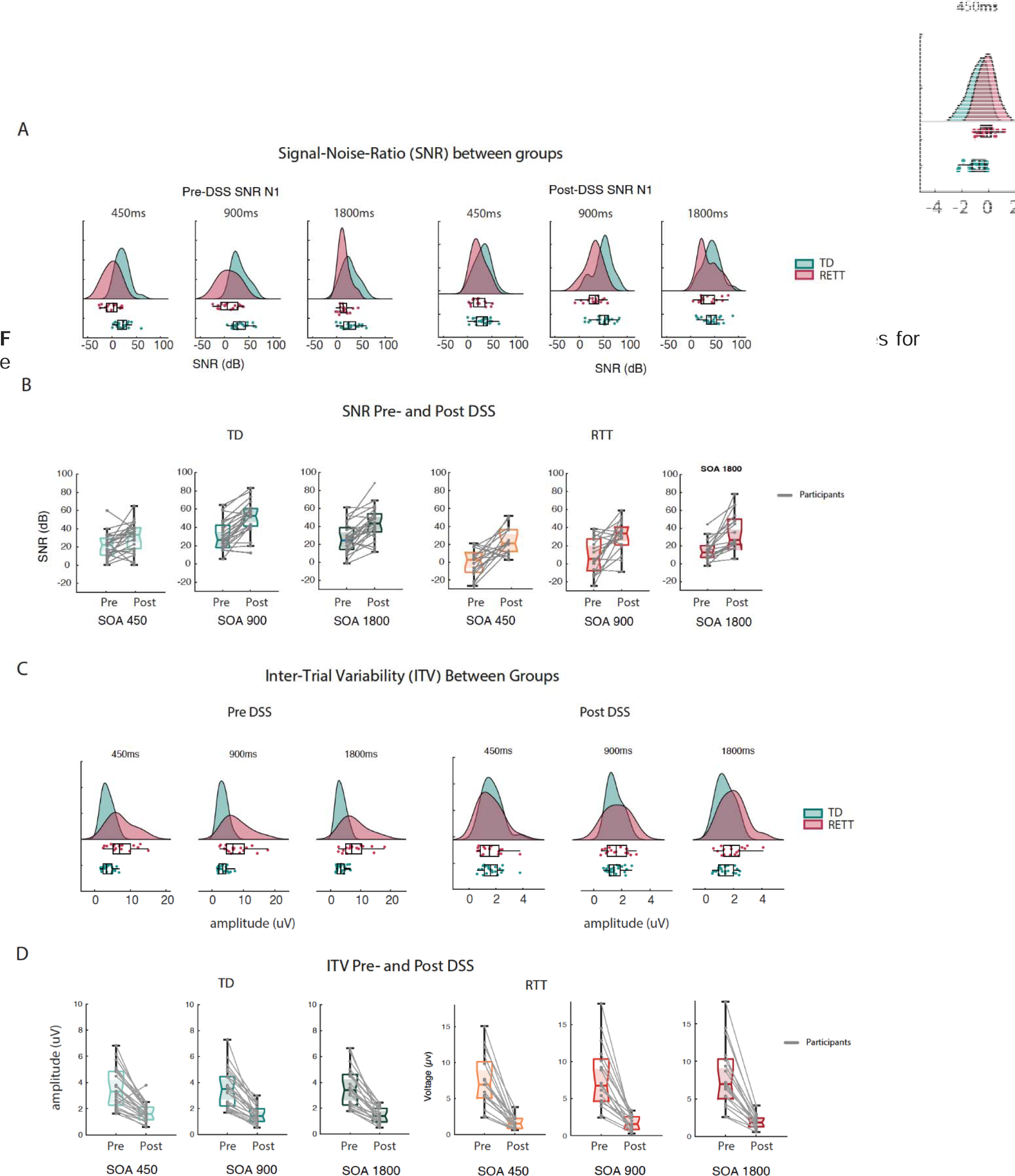
AEP values at N1 (200-300ms) across groups and SOA conditions. A. AEP values for each group and condition, pre- and post-DSS

*Linear-mixed effect (LME)* modeling on the N2 amplitude values revealed a significant main effect of Group (*Estimate* = 0.81, *p* = *p* 0.005, *t*_(1,241)_ = 2.81); *SE* = 0.28), DSS (*Estimate* = 0.58, *p* = 0.001, *t*_(1,241)_ = 3.21, *SE* 0.18), and Condition (*Estimate* = 0.13, *p* = 0.0004, *t*_(1,241)_ = 3.57, *SE* = 0.03). In addition, the Group*DSS interaction was significant (*Estimate* = −0.29, *p* = 0.015, *t*_(1,241)_ = −2.43, *SE* = 0.12), reflecting the reduced group difference for the DSS transformed data.

### Signal-to-Noise Ratio (SNR)

Comparisons of SNR between RTT and TD were conducted both pre- and post-DSS (Figure 5A and 5B and Table S3). Pre-DSS, SNR in RTT (*Mean ± sem = 7.75 ± 0.9dB*) was significantly lower than in TD (*Mean ± sem = 26.13 ± 0.64 dB; Wilcoxon rank test statistic = 5507; p < 0.001*). Post-DSS, SNR increased for the RTT group (*Mean ± sem = 30.06 ± 7.29 dB*) but remained significantly lower than TD (*Mean ± sem = 41.4 ± 8.45 dB; Wilcoxon rank test statistic = 5094; p = 0.0012*).

**Figure 5:** Pre- and Post DSS measures of signal-noise ratio (SNR) and inter-trial variability (ITV) for for RTT and TD in ERP peak N1. A. SNR: Between group comparison for each condition, pre- and post-DSS. B. SNR: Within group pre- and post-DSS comparison. C. ITV: Beween group comparison for each condition, pre- and post-DSS. **D.** ITV: Withing group pre- and post-DSS comparison.

Using an LME model with fixed effects of Group, DSS and Condition on SNR values, and participants as a random effect, a significant main effect of Group was revealed (*Estimate* = −14.85, *p* < 0.001, *t*_(1,242)_ = −4.63; *SE* = 3.21), in addition to an effect of DSS (*Estimate* = 18.2, *p* < 0.001, *t*_(1,242)_ = 9.6; *SE* = 1.89) and Condition (*Estimate* = 5.65, *p* < 0.001, *t*_(1,242)_ = 4.88− *SE* = 1.16). However, when including DSS*Group interaction in the model, neither the interaction, nor DSS as a main effect were significant (DSS: *Estimate* = 8.22, *p* = 0.15, *t*_(1,241)_ = 1.44; *SE* = 5.7; DSS*Group: *Estimate* = 7.04, *p* = 0.06, *t*_(1,242)_ = 1.85; *SE* = 3.8). Thus, DSS improved SNR for both groups of participants, but did not significantly reduce group differences in SNR. The estimated variance of the random intercepts was 24.9 *t*_(1,241)_ = 2.48; *p* = 0.013, indicating that there was significant variability among subjects in the intercept of the regression line. Thus, subjects as a random variable was a significant contributor to the overall variability.

### Inter-trial Variability (ITV)

To assess the degree of response stability in RTT participants, we calculated ITV on the N2 window for each participant (see Figure 5C, 5D). Pre-DSS, higher ITV was observed for RTT (*Mean ± sem = 7.76 ± 0.9µv*) than TD (*Mean ± sem = 3.5 ± 0.3µv*; *Wilcoxon rank test statistic*: *3152*; *p* = *1.66e-11*). Post-DSS, this ITV difference was no longer detected (*RTT: Mean ± sem = 1.75 ± 0.21; TD: Mean ± sem = 1.54 ± 0.13; Wilcoxon rank test statistic = 3152; p = 0.23)* (see Figure 5C and 5D and Table S3). Again, an LME model was implemented to analyze the ITV data with Group, SOA Conditions and DSS as fixed factors, while participants was a random factor. This analysis revealed a significant main effect of Group (*Estimate* = 8.2, *p* < 0.001, *t*_(1,242)_ = 11.08−; *SE* 0.74) and DSS (*Estimate* = 1.98, *p* < 0.001, *t*_1,241_^2^ = 3.68; *SE* = 0.53), but not for Condition (*Estimate* = 0.06; *t*_(1,241)_ = 0.55, *p* = 0.58, *SE* = 0.11). The Group*DSS interaction was significant (*Estimate* = −3.99; *t*_(1,241)_ = −11.12, *p* < 0.001, *SE* = 0.36), reflecting the decrease in group differences in ITV following the DSS procedure. The model included a random intercept for each individual to account for the correlation among the observations from the same subject. The estimated variance of the random intercepts was -2.75 (*p = 0.015*), indicating that there was significant variability among subjects in the intercept of the regression line.

### Inter-trial Phase Coherence (ITPC)

ITPC analysis was performed to measure the level of consistency of phase angle between single trials for each participant in each group, for each of the conditions. The ITPC data, illustrated in Figure 6, show an overall reduction in ITPC for the RTT compared to the TD group, and ITPC was higher overall for the data post-DSS. Figure 6A shows the ITPC dynamics along the 2-sec epochs, for each time point and frequency sample (1-25Hz). As seen in Figure 6A, lower ITPC for RTT is seen across conditions around times of stimulus onset. To assess the degree of ITPC, we calculated ITPC on the N2 window for each participant (see Figure 6B-C). Pre-DSS, higher ITPC was observed for TD (*Mean ± sem = 0.14 ± 0.002*) than RTT (*Mean ± sem = 0.103 ± 0.002*; *Wilcoxon rank test statistic*: *5207*; *p* = *1.37e-04*). Post-DSS, this ITPC difference became stronger (*TD: Mean ± sem = 0.26 ± 0.15; RTT: Mean ± sem = 0.15 ± 0.03; Wilcoxon rank test statistic = 5506; p = 8.82e-08)* (see Figure 6B and 6C for a group comparison, and DSS comparison, respectively).

**Figure 6:**
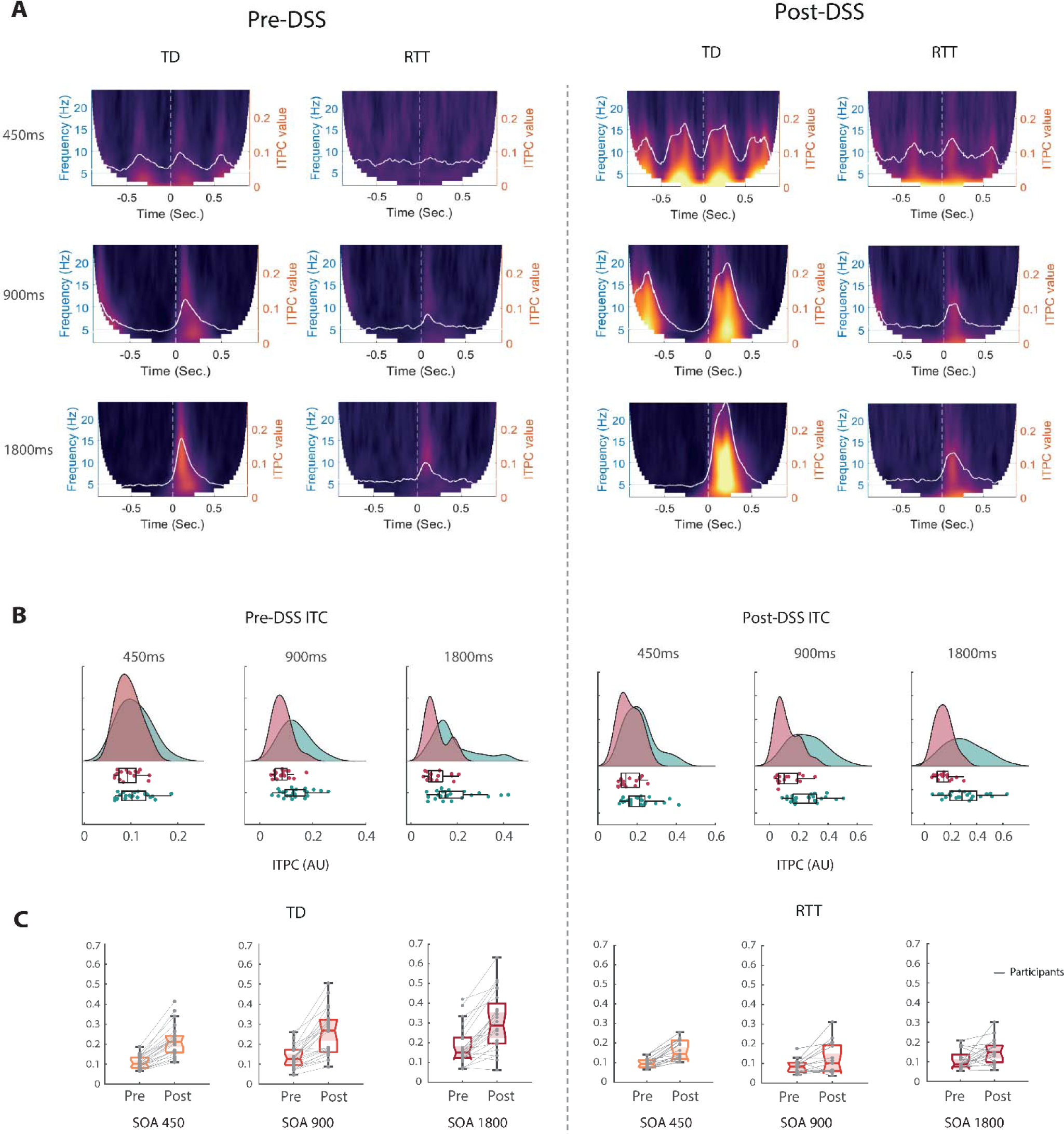
Inter-Trial phase coherence (ITPC) for each group and condition pre- and post-DSS. A. time-frequency ITPC plots show a reduction of coherence values in the RTT group across conditions, following stimulus onset. The average ITPC across frequencies is overlaid in white, on top of each plot. B. Raincloud plots of ITPC peak values derived from each participants (as seen in white in 4A), for each condition, pre- and post DSS. C. Same ITPC values shown in 4B, but now plotted within group for each condition, and compared between pre- and post DSS.

*A linear-mixed effect (LME)* model on the peak ITPC after stimulus onset revealed a significant main effect of Group (*Estimate* = −0.07, *p* = 7.96*e* − 05, *t*_(1,242)_ = −4.01); *SE* 002,), DSS (*Estimate* = 0.008, *p* = 3.96*e* −23, *t*_(1,242)_ = 11.01, *SE* = 0.008), and Condition (*Estimate* = 0.024, *p* = 9.8*e* −07, *t*_(1,242)_ = 5.02, *SE* = 0.005). When adding an interaction term of Group*DSS to the model, the interaction was significant (*Estimate* = −0.07, *p* = 6.99*e* − 06, *t*_(1,241)_ = −2.43, *SE* = 0.015), while the Group main effect was not (*Estimate* = 0.03, *p* = 0.3, *p* = 0.3, *t*_(1,241)_ = −2.43, *SE* = 0.015), reflecting that most of the variance in the group effect above was driven by one of the DSS conditions.

## DISCUSSION

Event-related potential recordings provide a simple, highly portable and relatively inexpensive means of directly and objectively recording neural processing outcomes from human subjects, even in patient populations where task compliance or the following of instructions is compromised or infeasible. Considerable work has now shown that measures of auditory evoked potentials (AEPs) are highly disordered in Rett syndrome (Foxe, Burke et al. 2016) (Key, Jones et al. 2019) (Brima, Molholm et al. 2019, Saby, Benke et al. 2021) (Peters, Katzenstein et al. 2017) (Badr, Witt-Engerstrom et al. 1987), and that these measures are correlated with clinical measures of disease severity (Sysoeva, Molholm et al. 2020). As a direct measure of neural function, therefore, these AEP measures hold much promise as neuromarkers against which the effectiveness of therapeutic interventions could be measured. It is a reasonable proposition that such measures are much closer to the site of action in pharmacological or gene therapy interventions and might be expected to show treatment-related changes much sooner than clinical observational measures that rely on changes in symptomatology or behavioral outcomes. These latter changes would be expected to emerge over relatively long timeframes, secondary to improvements in neural functioning. However, standard AEP/ERP signal processing techniques, which typically involve the averaging of multiple responses across trials, introduce significant risk of obscuring variability across individual neural responses, and could lead to overestimation of the extent of processing deficit that is actually present in a given individual. Here, the fact that there were three AEP conditions (i.e. three different inter-stimulus intervals were used in separate experimental blocks) allowed for a within-subject comparison across these conditions. What becomes clear upon simple visual inspection is that highly anomalous component morphology is common in RTT (see Figures 1 and 2), such that if an experimenter were to observe just one of these averaged AEPs for a given RTT participant, the presence of a response might be questionable. However, it is also clear from visual inspection that such anomalous morphologies are generally consistent across all three conditions – that is, similar appearing AEP responses are evident in most RTT participants. This is in contrast to the TD participants where for the most part, typical AEP morphology is observed. Put another way, in TD participants, there is a “central tendency” whereas in RTT participants, there is a tendency towards highly individualized responses. As mentioned in the Introduction, this has significant impact on group-averaged comparisons. Whereas the central tendency of the AEP in TDs will lead to a robust group-averaged waveform, the highly variable individual morphologies expressed in RTT will, by definition, lead to a weak group-averaged estimation. The highly idiosyncratic processing within the RTT group likely reflects disruption of typical processing along the auditory cortical processing hierarchy that does not manifest across patients in a stereotyped way.

Here, we set out to better understand this potential response variability by deploying a set of signal-processing tools at the single trial level, including denoising source separation, inter-trail variability estimation, inter-trial phase coherence measures, and estimates of signal-to-noise ratios. First, using the standard canonical component-based ERP analysis techniques, the current work replicated previous outcomes of strikingly atypical group level AEPs in individuals with RTT, which were evident at each of the three stimulation rates used (Figures 1 and 2) (Brima, Molholm et al. 2019), differences that were manifest as substantially reduced AEP component amplitudes in RTT versus TD (Sysoeva, Molholm et al. 2020) (Saby, Benke et al. 2021). In turn, taking advantage of the large number of trials per condition that were recorded in this study, we assessed inter-trial response variability to the auditory stimuli at the individual participant level. Across all metrics: signal-to-noise ratio (SNR), inter-trial variability (ITV) (see Figure 5), and inter-trial phase coherence (ITPC: Figure 6), significantly higher levels of response variability were observed in RTT compared to the TD participants.

Another possible confounding factor when comparing RTT, or indeed any clincial population, to TD controls using EEG meaures is that neural repsonses might be obscured by excessive non-neural noise (e.g. movement artifacts, muscle tension or flexion noise, excessive eye-movements or blinking, bruxism, etc.). To mitigate such influences, we also applied the denoising source separation (DSS) algorithm, a joint-decorrelation technique that suppresses the most prominent non-neural noise sources, and preserves the activity of interest (de Cheveigné and Parra 2014). The previous set of analyses were then repeated using these DSS accentuated signals. Clear improvements in the signal post-DSS were observed for both groups, but especially so for the RTT group, such that large between-groups differences in the amplitude of the N2 AEP component and measures of inter-trial variability (ITV) that were observed prior to denoising, were no longer statistically detectable following application of the technique. However, post-DSS measures of SNR remained significantly lower in the RTT group, and differences between RTT and TD in ITPC were even more robust following DSS. Taken together, these analyses make clear that non-neural sources of noise very likely contribute to overestimation of the extent of AEP deficits in RTT but that clear deficits remain detectable following denoising that minimizes the contribution of non-neural noise to response estimates. This suggests that application of DSS should likely be a facet of any signal-processing pipeline designed to test neural information processing in individuals with rare diseases like RTT.

The SNR calculations pre-versus post-denoising are highly instructive in this regard, demonstrating clear and rather dramatic effects of applying DSS. In the case of the RTT group, SNR across conditions increased from 7.8 to 30.1, representing a massive 3.9-fold increase in signal estimation. SNR did also increase in the TD group, but by a more modest amount (26.1 to 41.4), a 1.6-fold increase. While SNR in RTT remained significantly lower than that found in TD, it is clear that pre-denoising, this difference was substantially overestimated and suggested a much greater deficit than is likely the case. Non-evoked potential noise is therefore a major source of potentially confounding variance in inter-group comparisons concerning RTT individuals and should be a consideration in all studies assessing differences between rare-disease clinical groups and neurotypical control populations. Similarly, AEP peak voltage variability, assessed by ITV, improved post-DSS in both groups, but more so in RTT (a 4.5-fold decrease, from 7.76 to 1.75µv) than TD (a 2.3-fold decrease, from 3.50 to 1.54 µv). This time, group differences in ITV were, in fact, no longer statistically detectable post-DSS. Lastly, in the case of ITPC, DSS also significantly improved these estimates in both populations. In the TD group, ITPC estimate pre-DSS was 0.14 but this improved to 0.26 following denoising, whereas in the RTT group the improvement was more modest (0.10 to 0.15). For both pre- and post-DSS estimates, the difference between TD and RTT participants was statistically robust.

Thus, while denoising substantially improves SNR in RTT and leads to lower estimates of inter-trial variability both in terms of response amplitudes and phase, substantial deficits remained in the RTT group in SNR and ITPC measures of response variability, whereas this was not the case for ITV. In summary, while previous results showed robust evoked-response atypicalities using the standard component-based ERP approach, the present work suggests that the addition of measures that assess response variability can add significant insight into putative dysfunction and may well provide more sensitive biomarkers for assessment of treatment effects on neural function. That ITPC is found to be significantly lower in RTT, even post-DSS, provides at least partial support for a neural unreliability account of auditory processing deficits in this population, although lower SNR estimates and idiosyncratic temporal evolution of the AEP also suggest that sensory processing is both attenuated and temporally disrupted, and that the differences between RTT and TD are not wholly accounted for by “unreliable” responsivity.

### Study Limitations

Auditory responses continue to mature with typical development (Bishop, Anderson et al. 2011, Brandwein, Foxe et al. 2011, Brandwein, Foxe et al. 2013), and as such, our relatively wide participant age range (7 to 22 years of age) is a limiting factor. Furthermore, the number of usable RTT data sets was reduced from 25 to 17 due to excessively noisy EEG data and an insufficient number of accepted trials per condition. It will be key to develop better methods to capture adequate EEG data in these difficult-to-test populations, as a 68% success rate will not be adequate if such measures are to be fully useful as outcomes in clinical trials. It is also the case that the limited RTT sample precludes the possibility to meaningfully examine mutation subtype in this cohort due to the lack of sufficient power. Neither were we able to consider potential differences as a function of classic versus atypical Rett phenotype. Both of these distinctions will be of great interest as this work progresses.

### Conclusions

This study deployed in-depth analysis of auditory evoked response variability to assess the contribution of the degree of response variability (unreliability) to altered auditory processing in RTT. We replicated previous outcomes of atypical AEP morphologies and significantly reduced AEP amplitudes in Rett Syndrome using standard component-based ERP analysis. Using metrics that specifically measured neuronal variability, we observed substantially increased inter-trial variability, lower signal-to-noise ratios, and reduced inter-trial phase coherence in the auditory responses of RTT participants, providing strong support for a “neural unreliability” account in this population. However, deployment of denoising source separation (DSS) techniques painted a somewhat different picture, making it clear that non-neural sources of noise are a likely contributor to overestimation of the extent of auditory processing deficits in this population. Post-DSS, ITV measures were substantially reduced, so much so that pre-DSS ITV differences between RTT and TD populations were no longer detected. In the case of SNR and ITPC, DSS substantially improved these estimates in the RTT population, but robust differences between RTT and TD were still fully evident. This work strongly suggests that employing DSS techniques will provide much better estimates of veridical sensory-perceptual processing abilities in rare disease populations such as RTT, or in any other population where a high degree of non-neural noise and high inter-individual variability are expected to be major contributors.

## Data Availability

All data produced in the present study are available upon reasonable request to the authors

## Abbreviations List

AEP: Auditory Evoked Potentials
ANOVA: Analysis of Variance
DSS: Denoising Source Separation
EEG: electroencephalogram
ERP: Event-Related Potential
ITPC: Inter-trial Phase Coherence
ITV: Inter-trial Variability
LME: Linear Mixed Effects
MMN: Mismatch Negativity
RSSS: Rett Syndrome Severity Scale
RTT: Rett Syndrome
SNR: Signal to Noise Ratio
SOA: Stimulus Onset Asynchronies
TD: Typically Developed

## DECLARATIONS

### Ethics approval and consent to participate

All aspects of the research conformed to the tenets outlined in the Declaration of Helsinki, with the exception that this study was not preregistered. The institutional review boards of the University of Rochester and of the Albert Einstein College of Medicine approved this study. Written informed consent was obtained from parents or legal guardians. Where possible, informed assent from the participants was also obtained.

### Consent for publication

Not applicable

### Author contributions

JJF and SM conceived the study and designed the original experiment. TB and AD recruited and phenotyped the participants. TB collected the data. TB, SB, JSB and KDP analyzed the data and created the illustrations. JSB and KDP designed and consulted on the single-trial analysis. TB wrote the first draft of the paper, in close collaboration with SB and KDP. JJF, SM and EGF provided substantial editorial input and writing on subsequent drafts. EGF, KDP, JSB and AD provided critical theoretical input and manuscript revision. All authors read the final draft and provided critical input.

## Acknowledgments

The authors acknowledge the participation of Mr. Douwe Horsthuis and Mr. Emmett Olan Foxe during the data collection phase of this project. We also thank the volunteers, the patients, and their families, who graciously gave of their time with great patience to allow for the successful completion of this study.

## Funding

This work was supported by a grant from the U.S. National Institute on Deafness and Other Communication Disorders (NIDCD R21 DC012447 to JJF) and with generous funding from The Rett Syndrome Research Trust. Recordings and participant phenotyping were accomplished through cores of the University of Rochester Intellectual and Developmental Disabilities Research Center (UR-IDDRC) which is supported by a center grant from the Eunice Kennedy Shriver National Institute of Child Health and Human Development (P50 HD103536 – to JJF) and the Rose F. Kennedy Intellectual and Developmental Disabilities Research Center (RFK-IDDRC), supported by the Eunice Kennedy Shriver National Institute of Child Health and Human Development (P50 HD105352 – to SM).

## Conflicts of interest

The authors declare no financial or other competing interests that are pertinent to the results of this study.

## Availability of data and material

The datasets used and/or analyzed during the current study are available from the corresponding author on reasonable request.

## Supplementary figures

**Figure S1:**
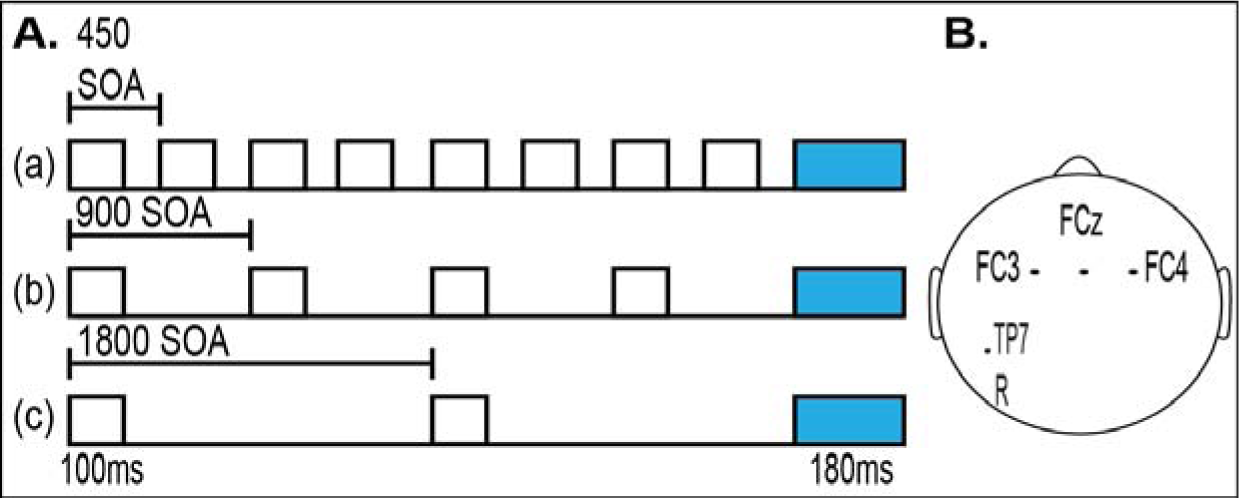
Oddball paradigm design: A. 3 experimental conditions with varied stimulus presentation intervals (SOA): (a) 450 ms intervals between stimuli, (b) 900 ms between stimuli and (c) 1800 ms between stimuli. B. Shows a map of the electrode site of interest (FC3, FCz, and FC4) and reference electrode R (TP7).

**Table S1.** Clinical demographics of all enrolled participants with Rett syndrome. The shaded area depicts excluded participants. RSSS = Rett Syndrome Severity Score; Seizure OS = Seizure on set; Age of R = Age of regression in months (mos.); N/A = Not Available.

| Subject | Age | Mutations | (RSSS) | Seizures | Seizure OS | Ambulatory | Medications | Age of R |
| --- | --- | --- | --- | --- | --- | --- | --- | --- |
| 1 | 6-9 | R255X | 13 | yes | 48 mos | no | Trazadone, Depakote | N/A |
| 2 | 6-9 | R133C | 8 | yes | 72 mos | yes | Lovastatin, Topomax, Abilify, Lexapro, Depakene, Nexium | 21 mos |
| 3 | 10-14 | R306C | 7 | yes | 168 mos | yes | Lovastatin, Ambien, Trazodone | 50-60 mos |
| 5 | 10-14 | C964C | 5 | no | no | yes | Trazadone | 156 mos |
| 7 | 6-9 | deletion | 9 | no | no | yes | Lovastatin | 11 mos |
| 8 | 10-14 | R255X | 6 | no | no | yes | Lovastatin | 12 mos |
| 9 | 15-19 | R270X | 14 | N/A | N/A | no | none | N/A |
| 13 | 10-14 | deletion | 14 | yes | 30 mos | no | Depakene, Lexapro, Lactulose | N/A |
| 14 | 10-14 | Q170x mutation at nucleotide c508 | 10 | N/A | N/A | yes | N/A | 18 mos |
| 16 | 10-14 | deletion | 12 | no | 72 mos | yes | Risperdol and Necon | 144 mos |
| 17 | 20-24 | R133C | 12 | yes | 108 mos | yes | Depakote, Carintor, Artane, Copaxone | 17 mos |
| 18 | 10-14 | deletion | 11 | N/A | N/A | no | Depakene | N/A |
| 19 | 15-19 | R270X | 13 | no | N/A | no | N/A | N/A |
| 21 | 15-19 | 418ins4 | 15 | yes | N/A | no | Diastat, Depakene, Carnitor | N/A |
| 22 | 20-24 | T158M | 11 | yes | 72 mos | yes | Lexapro, Lamictal | 15-18 mos |
| 23 | 6-9 | Large deletion | 12 | yes | 60 mos | no | Valproic acid | N/A |
| 24 | 6-9 | Deletion exon 3&4 | 9 | yes | 36 mos | no | Keppra, deplane | N/A |
| 25 | 10-14 | T158M | 12 | no | N/A | no | Nanadol, prevacid | 24 mos |
| 4 | 15-19 | T158M | 11 | yes | 120 mos | yes | Copaxone, Topomax, Lovastatin, | 10 mos |
| 6 | 6-9 | R294X | 7 | no | no | yes | Lexapro, Lovastatin, Prilosec | 17 mos |
| 10 | 10-14 | R306C | 9 | no | no | yes | Lexapro | 18 mos |
| 11 | 15-19 | R306C | 10 | no | no | yes | Lexapro, Copaxone, Trazodone, | N/A |
| 12 | 6-9 | deletion | 7 | yes | 24 mos | no | Depakene, Lactulose, Lovastatin, | 30 mos |
| 15 | 10-14 | T158M | 10 | N/A | N/A | no | N/A | N/A |
| 20 | 6-9 | R133C | 10 | yes | 42 mos | yes | Abilify, Lexapro, Desyrel | 24 mos |

**Table S2.** Number of accepted standard trials and grand average AEP values and corresponding standard deviations (±SD) included in the analysis across conditions in typically developing controls and in participants with Rett syndrome.

|  | TD (N=24)<br>Mean Age: 12.3 ± 4.7 |  |  | RTT (N=17)<br>Mean Age: 12.6 ± 4.8 |  |  |
| --- | --- | --- | --- | --- | --- | --- |
|  | 450 SOA | 900 SOA | 1800 SOA | 450 SOA | 900 SOA | 1800 SOA |
| Average Accepted Std. trials ± SD | 412.5 ± 153 | 718.5 ± 96 | 720 ± 81 | 239.1 ± 81 | 507.6 ± 153 | 460.2 ± 96 |
| Accepted Overall | 1851 |  |  | 1207 |  |  |
| Grand average AEP values ± SD | 1.18 ± 0.85 | 0.79 ± 1.01 | 0.61 ± 1.17 | 0.64 ± 0.90 | 0.25 ± 0.99 | -0.13 ± 1.07 |

**Table S3.** Mean ± SEM for SNR and ITV, Pre and Post–DSS, for each condition in typically developing (TD) controls and in participants with Rett syndrome (RTT).

|  |  | TD (N=24) |  |  | RTT (N=17) |  |  |
| --- | --- | --- | --- | --- | --- | --- | --- |
|  |  | 450 SOA | 900 SOA | 1800 SOA | 450 SOA | 900 SOA | 1800 SOA |
| SNR | Pre-DSS Mean ± SEM | 21.09 ± 2.77 | 30 ± 3.25 | 26.55 ± 3.4 | -0.81 ± 3.72 | 9.29 ± 4.76 | 14.88 ± 2.86 |
|  | Mean ± SEM Across Conditions | 26.13 ± 0.64 |  |  | 7.75 ± 0.91 |  |  |
|  | Post-DSS ± SEM | 30.98 ± 3.25 | 49.75 ± 3.86 | 43.46 ± 3.69 | 23.13 ± 3.56 | 30.29 ± 4.26 | 36.76 ± 5.02 |
|  | Mean ± SEM Across Conditions | 41.4 ± 8.45 |  |  | 30.06 ± 7.29 |  |  |
| ITV | Pre-DSS Mean ± SEM (µv) | 3.57 ± 0.3 | 3.55 ± 0.3 | 3.54 ± 0.29 | 7.35 ± 0.88 | 7.93 ± 1.03 | 8.01 ± 1.05 |
|  | Mean ± SEM (µv) Across Conditions | 3.55 ± 0.29 |  |  | 7.76 ± 0.97 |  |  |
|  | Post-DSS ± SEM (µv) | 1.68 ± 0.14 | 1.52 ± 0.13 | 1.41 ± 0.12 | 1.59 ± 0.21 | 1.72 ± 0.23 | 1.95 ± 0.21 |
|  | Mean ± SEM (µv) Across Conditions | 1.54 ± 0.13 |  |  | 1.75 ± 0.21 |  |  |

## Notes

### Competing Interest Statement

The authors have declared no competing interest.

